# Identification of EPHB4 variants in dilated cardiomyopathy patients

**DOI:** 10.1101/2022.12.16.22283442

**Authors:** Guillermo Luxán, Marion Muhly-Reinholz, Simone F. Glaser, Johannes Trebing, Christoph Reich, Jan Haas, Farbod Sedaghat-Hamedani, Benjamin Meder, Stefanie Dimmeler

**Author notes:** **Corresponding author** Guillermo Luxán, Goethe University Frankfurt, Theodor Stern Kai 7, 60590 Frankfurt; Germany.

## Abstract

Cardiac homeostasis relies on the appropriate provision of nutrients to the working myocardium. EPHB4 is required for the maintenance of vascular integrity and correct fatty acid transport uptake in the heart via regulating the caveolar trafficking of the fatty acid receptor CD36. In the mouse, endothelial specific loss-of-function of the receptor EphB4, or its ligand ephrin-B2, induces Dilated Cardiomyopathy (DCM) like defects. Now, we have identified six new *EPHB4* variants with deleterious potential in a cohort of 573 DCM patients. Similar to the EphB4 mutant mice, *EPHB4* variants carrying patients show an altered expression pattern of CD36 and CAV1 in the heart. For the first time, our data identifies EPHB4 mutations in DCM patients. This observation supports the notion that the Eph-ephrin signalling pathway, and in particular the receptor EPHB4, plays a role in the development of DCM in human patients.

## Introduction

Dilated cardiomyopathy (DCM) is one the most common type of heart disease with an estimated prevalence of 1:2500 in the general population^1^. DCM patients are characterized by the dilation of the left ventricle and contractile dysfunction^2^. The definition of DCM excludes ischemic heart disease, although ischemic events as induced by myocardial infarction can lead to similar remodelling processes and contractile dysfunction resembling DCM^3^.

The most common causes of DCM underlie inflammation, nutritive-toxic influence, or metabolic disorders^4^ but mutations on genes encoding cytoskeletal, sarcomere or nuclear envelope proteins account for about 35% of DCM cases^2^. Increasing evidence shows that heart disease is associated with microvascular dysfunction^5^. Endothelial cell CD36 is required for the uptake of circulating fatty acids into the cardiac muscle^6^. In the mouse, interference with endothelial nutrient transport resulted in reduced cardiac function^7^. Specifically, endothelial specific deletion of Ephrin type-B receptor 4 (EPHB4), or its ligand ephrin-B2, induces cardiac remodelling and a phenotype that mimics human DCM features and development^8^. Mechanistically, EPHB4 deficient endothelial cells are characterized by compromised caveolar function and reduced Caveolin 1 (CAV1) phosphorylation. Caveolae are required for the correct membrane translocation of the fatty acid translocase FAT/CD36^9^ and fatty acids are used by cardiomyocytes to obtain about 50% to 70% of their energy^10^.

In the present study, we identify six new EPHB4 mutations in a cohort of DCM patients. These patients, similar to the EPHB4 mutant mice, show dysregulated expression of CAV1 and CD36 although in the studied patients, this was observed in the cardiomyocytes. Our results confirm a crucial role of the Eph-ephrin signalling pathway in DCM.

## Methods

### Human samples

All participants in the DZHK-TORCH and in the registry of the Institute for Cardiomyopathies Heidelberg registry have given written informed consent and the study was approved by the ethic committees of the participating study centres. A peripheral blood sample was collected from each participant. SOP Quality Training and monitoring were performed regularly as described^11^.

### Immunohistochemistry

Human sample biopsies were fixed with 4% PFA and embedded in paraffin. Rabbit anti-CAV1 (1:100, 3238S, Cell Signaling, RRID:AB_2072166), mouse anti-CD36 (1:50, C114979, LSBio), and Biotinylated Ulex (1:50, B-1065, Vector, RRID:AB_2336766) were applied overnight, followed by a 1 hour incubation at room temperature with donkey anti rabbit Alexa Fluor 555 (1:200, A31572, Invitrogen, RRID:AB_162543), donkey anti mouse Alexa Fluor 488 (1:200, A21202, Invitrogen, RRID:AB_141607), and Streptavidin Alexa Fluor 647 (1:200, S32357, Invitrogen). Researchers were blind to patient genotype and status for immunohistochemistry and analysis. 6 EPHB4 variant carrying DCM patients, 6 DCM patients with wild type EPHB4 and one healthy control biopsy.

### Single-nucleus-RNA-sequencing

The single-nucleus-RNA-sequencing data set generated in the paper by Nicin et al.^12^ was used to explore *EPHB4* expression in human cardiac cells.

## Results

To assess if EPHB4 mutations might play a role in human heart failure, we analysed the EPHB4 sequences of in total 573 DCM patients, which were either enrolled in the **T**ranslati**O**nal **R**egistry for **C**ardiomyopat**H**ies-Plus (TORCH-Plus-DZHK21)^13^, or recruited at the Institute for Cardiomyopathies Heidelberg (ICH). In six patients, of which histology was also available, we found six new EPHB4 variants (**Table 1**). Three of the newly identified variants are located in the extracellular domain of EPHB4, two of them in the ligand binding domain of the protein and another one on the cysteine rich domain before the two fibronectin type III domains. The other three variants are located in the intracellular part of the EPHB4 protein, in the tyrosine kinase domain (**Figure 1A**). In four of these patients we did not detect any mutation in genes previously related to DCM while in the other two patients we could also detect a truncating *RMB20* or *TTN* variants. The Combined Annotation Dependent Depletion (CADD) score of these variants (21,64 [IQR 19,41-23,50]) indicates that the variants found in this study have a high potential to be deleterious as CADD scores >20 indicate the top 1% of predictably deleterious variants. In order to understand whether these new EPHB4 variants could be implicated in the disease, we analysed DCM patients carrying the heterozygous EPHB4 variant and DCM patients with no such variant. We furthermore compared the results with a biopsy obtained from a healthy donor. The basal characteristics of the patients can be found on **Table 2**.

**Table 1.** Overview on EPHB4 variants. Overview on exonic EPHB4 variants found in whole genome and whole exome sequencing data, respectively. Together, 573 individuals with DCM have been analysed. The frequencies within DCM cases and population controls (gnomAD) are given. CADD_phred scores >20 indicate the top 1% of predictably deleterious variants.

| Gene | ENSG_ID | ENST_ID | cDNA | AAchange | rs_dbSNP | CADD_phred | gnomAD_exomes_controls_NFE_AF | DCM allele frequency | DC M1 | DC M2 | DC M3 | DC M4 | DC M5 | DC M6 |
| --- | --- | --- | --- | --- | --- | --- | --- | --- | --- | --- | --- | --- | --- | --- |
| EPH B4 | ENSP00000350896.3 | ENST00000358173.8 | c.1905G>T | p.Lys635Asn | rs374676243 | 24.1 | 0.00002338415 | 0.0018 | 1 | 0 | 0 | 0 | 0 | 0 |
| EPH B4 | ENSP00000350896.3 | ENST00000358173.8 | c.610C>G | p.Leu204Val | NA | 22.9 | NA | 0.0018 | 1 | 0 | 0 | 0 | 0 | 0 |
| EPH B4 | ENSP00000350896.3 | ENST00000358173.8 | c.337G>A | p.Val113Ile | rs55866373 | 22.4 | 0.0003507623 | 0.0035 | 0 | 1 | 0 | 0 | 0 | 1 |
| EPH B4 | ENSP00000350896.3 | ENST00000358173.8 | c.2670G>T | p.Glu890Asp | rs35638378 | 17.22 | 0.008748404 | 0.0209 | 0 | 0 | 1 | 1 | 0 | 0 |
| EPH B4 | ENSP00000350896.3 | ENST00000358173.8 | c.2051_2052del | p.Thr684fs | NA | NA | NA | 0.0018 | 0 | 0 | 1 | 0 | 0 | 0 |
| EPH B4 | ENSP00000350896.3 | ENST00000358173.8 | c.236C>T | p.Pro79Leu | rs542089930 | 21.6 | 0.0003295048 | 0.0052 | 0 | 0 | 0 | 0 | 1 | 0 |

Gene= gene name; ENSG\_ID=Ensemble gene ID; ENST\_ID=Ensemble transcript ID; cDNA=predicted cDNA change for the listed transcript; AAchange=predicted amino acid change for the listed transcript; rs\_dbSNP=dbSNP ID; CADD\_phred= CADD score prediction; gnomAD\_exomes\_controls\_NFE\_AF= gnomAD allele frequency in the exome sample of non-finnish european descent; DCM allele frequency= allele frequency of the variant in full cohort of 573 individuals; DCM1-6= genotype for the DCM patients where 0=homozygous reference and 1=heterozygous.

**Table 2.** Baseline characteristics of the patient cohort.

|  | DCM patients<br>(reference genome) | DCM patients<br>(EPHB4 variant) |
| --- | --- | --- |
| n | 6 | 6 |
| Age (years) | 60 [IQR 48-75] | 43 [IQR 29-56] |
| Sex (male/female) | 3/3 | 5/1 |
| NYHA class [I/II/III/IV (n)] | 1/0/3/1 | 0/3/2/0 |
| Hypertension [n (%)] | 5 (83.3) | 3 (50) |
| Hyperlipidemia [n (%)] | 3 (50) | 4 (66.6) |
| Diabetes [n (%)] | 3 (50) | 2 (33.3) |
| Smoking [n (%)] | 3 (50) | 3 (50) |
| Left ventricular EF (%) | 35.17 [IQR 20-48.5] | 30.20 [IQR 17.5-40.5] |
| ACEi or ATRB (%) | 6 (100) | 4 (83.3) |
| Beta-blocker (%) | 6 (100) | 5 (83.3) |
| Diuretic (%) | 4 (66.6) | 3 (50) |
| Aldosterone antagonist | 2 (33.3) | 3 (50) |
| Statin | 4 (66.6) | 4 (66.6) |

DCM – dilated cardiomyopathy
EF – ejection fraction
ACEi – angiotensin-converting enzyme inhibitor
ATRb - *angiotensin* II receptor *blocker*

**Figure 1.**
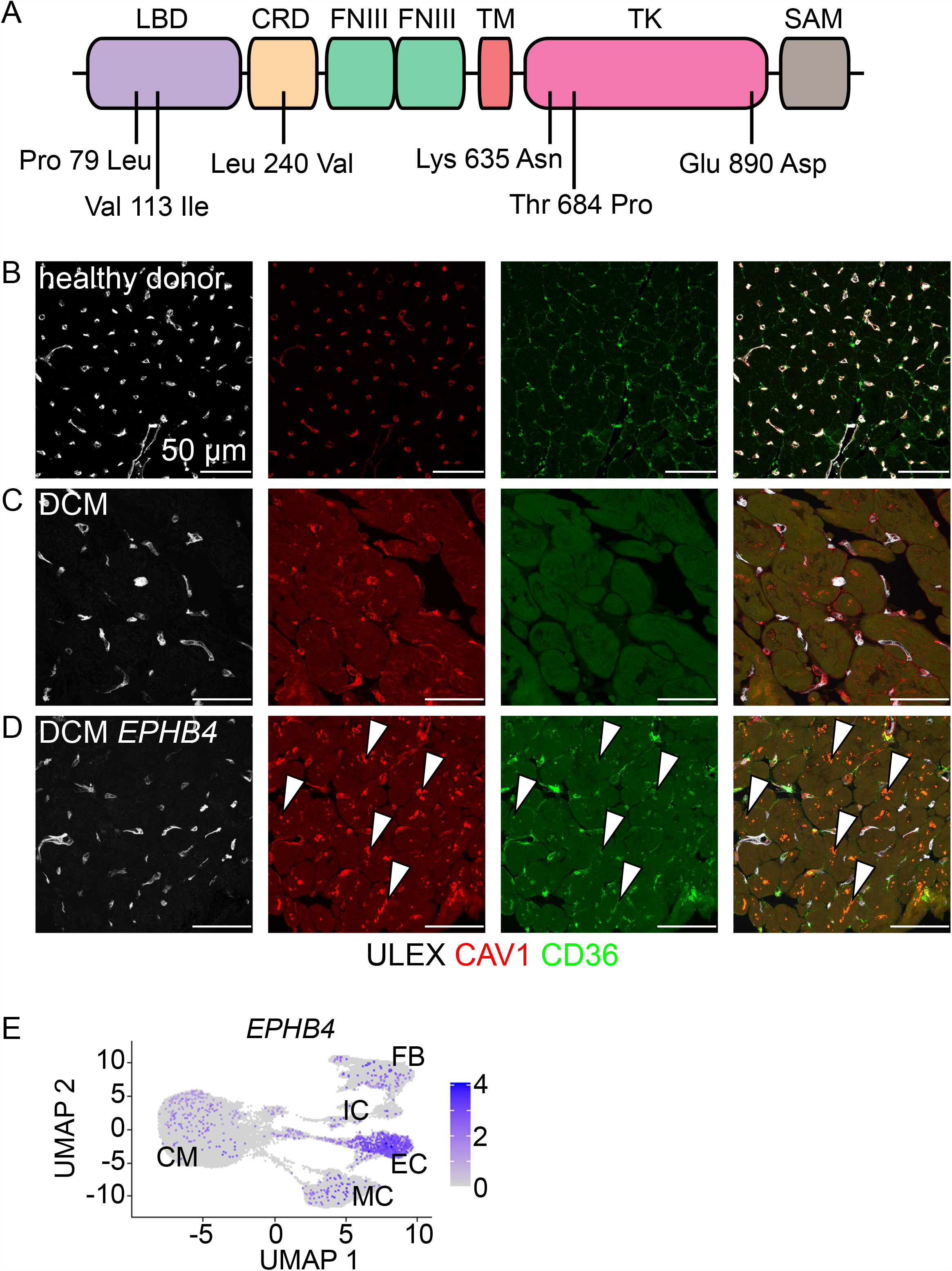
EPHB4 mutations in human DCM patients. (**A**) Scheme depicting the EPHB4 protein with the mutations indicated in the protein domains. (LBD, ligand binding domain; CRD, cysteine-rich domain; FNIII, fibronectin type III; TM, transmembrane domain; TK, tyrosine kinase; SAM, sterile alpha domain). (**B, C, D**) Immunohistochemistry of cardiac biopsies from healthy donor (**B**), DCM patients with reference genotype (**C**), and DCM patients carrying the *EPHB4* variant (**D**). CAV1 and CD36 expression is altered in both DCM conditions. CD36 expression is reduced in the *EPHB4* variant carrying patients and it colocalizes with CAV1 in the cardiomyocytes (**arrowheads**). (**E**) *EPHB4* expression in the human heart analysed by single-nuclei RNA sequencing. Cardiac cells are displayed in a UMAP plot. Although *EPHB4* is mainly expressed by endothelial cells, *EPHB4* is also expressed in some cardiomyocytes. Colour reflects expression levels in each nucleus. (CM, cardiomyocyte; FB, fibroblasts; IC, immune cell; EC, endothelial cell; MC, mural cell).

Histological analysis of cardiac biopsies obtained from these patients compared to biopsies from DCM patients with reference genotype and healthy donors revealed that the expression of CD36 and CAV1 is altered in DCM patients. CD36 is highly expressed in the capillaries and in the membrane of cardiomyocytes of healthy hearts, while CAV1 can be detected in the capillary endothelial cells (**Figure 1B**). In DCM patients which carry a wild type EPHB4 sequence, we noted that the expression of CD36 is abrogated and a disorganized expression of CAV1 can be observed in the cardiomyocytes (**Figure 1C**). In the *EPHB4* variant carriers, the expression of CD36 is reduced in the membrane of cardiomyocytes but colocalizes with CAV1 in accumulated vesicle structures in the cardiomyocytes, suggesting impaired CD36 caveolar transport (**Figure 1D, arrowheads**). Interestingly, and different to the data on mouse ^8^, the miss localization of CAV1 and CD36 was observed in cardiomyocytes.

Analysis of available human cardiac single-nuclei-RNA-sequencing^12^ revealed that *EPHB4* is also expressed in human cardiomyocytes (**Figure 1E**) suggesting that EPHB4 may control CD36 localization also in cardiomyocytes.

## Discussion

Our results suggest that the dysfunction of the EPHB4 receptor and its regulation of caveolae trafficking required for fatty acid uptake as a potential cause for DCM in human patients. Furthermore, our results stress the importance of the endothelial CD36 in the onset of cardiac disease as all DCM patients show a downregulation of CD36 in the endothelium and warrant a more detailed assessment of genes involved in vascular function^14^. Nevertheless, although more abundantly expressed in endothelial cells, CD36 is also expressed in cardiomyocytes^15^ and its loss in cardiomyocytes has already been related with the progression of heart failure^16^. There are different molecular mechanisms that can regulate the expression and cellular localization of CD36. CD36 expression is upregulated by the nuclear hormone transcription factor PPAR-γ and cytokines like CSF and IL4. In the other hand, LPS and dexamethasone downregulate its expression In microvascular endothelial cells, CD36 is downregulated by lysophosphatidic acid. Glycosylation also impacts CD36 expression levels, as does lipid acylation of both the N-terminal and C-terminal intracellular domains (reviewed in^17^). We hypothesize that mutations or alteration on these pathways in the heart, similar to EPHB4, may also cause DCM and should be further studied.

Cardiomyocytes are the cells responsible for generating contractile force in the heart and were traditionally at the center of interest in genetic studies. However, only about a third of DCM cases can be explained by mutations on structural cardiomyocyte genes^2^. This study suggests that mutations in genes crucial for cell to cell signalling may contribute to DCM. Attempts to use single-cell- and single-nucleus-RNA-sequencing of cardiac tissue of patients^18^ to identify potential genes and pathways in the other cell types just started and certainly will enrich our understanding of the multi-cellular communication that is necessary to maintain cardiac health.

Our analysis identified several variants in EPHB4 in a cohort of DCM patients. According to the CADD score prediction, all these variants have a deleterious potential. During Eph-ephrin signalling, the binding of the ligand induces Eph receptor heterotetramers to initiate the signalling via Eph–Eph cis interactions^19^. Thus, variant EPHB4 molecules could have a dominant negative effect on these heterotetramers explaining why the presence of one variant copy in the DCM patients of our cohort would be sufficient to abrogate the function of the protein. The histology analysis of the cardiac biopsies revealed that, independently of where in EPHB4 the variant was detected, the EPHB4 variant carrying patients show similar histological features with CD36 and CAV1 colocalizing in vesicular structures and are different to the other DCM patients with a wild type EPHB4. This observations includes the two patients that carry variants in other DCM related genes. Although we can’t discard the contribution of the variants in other genes in two of the patients, this study further supports, the importance of EPHB4 regulating CD36 caveolar trafficking to the membrane, whether this happens in endothelial cells or cardiomyocytes, maintaining cardiac homeostasis in humans and its implication on DCM. Recent studies suggest that Eph receptor genes, including *EPHB4*, were downregulated in hypertrophic human hearts^12^ suggesting that a transcriptional or post transcriptional down-regulation of Eph receptors might also contribute to acquired forms of heart failure. Interestingly, beyond its function in the cellular membrane, circulating EPHB4 is associated with poor prognosis in heart failure^20^.

Finally, this study not only confirms the crucial role of EPHB4 in the heart, but it also corroborates the importance of CD36 and CAV1 for the cardiac health, and has the potential to improve diagnosis and risk stratification tools for DCM. In addition, as other genes crucial for fatty acid transport may be involved in cardiac disease, this study may help identify new diagnostic or therapeutic targets.

## Data availability

The authors declare that the data underlying the findings of this study are available within the paper or are available upon request per

## Ethics approval and consent to participate

All participants in the DZHK-TORCH and in the registry of the Institute for Cardiomyopathies Heidelberg registry have given written informed consent and the study was approved by the ethic committees of the participating study centres.

## Author Contribution

GL and SD contributed to the conception and design of the research. GL analyzed data, acquired funding and managed the project. MMR, performed immunohistochemistry. SFG analyzed the single-nucleus-RNA-sequencing. JT, CR, JH, FSH and BM analyzed the databases and provided histology samples of DCM patients. GL and SD wrote the paper.

